# Genes, gut microbiome, and lungs: Untangling the web of Idiopathic Pulmonary Fibrosis

**DOI:** 10.1101/2025.10.30.25339139

**Authors:** Tamara Hernandez-Beeftink, Richard J. Allen, Olivia C. Leavy, Leah Cuthbertson, Philip L. Molyneaux, Louise V. Wain

## Abstract

Idiopathic pulmonary fibrosis (IPF) is a progressive lung disease with limited treatment options and poor survival. The airway microbiome plays a critical role in IPF, and there is emerging interest in the potential involvement of the gut microbiome through the gut-lung axis. Previous studies have reported genetic associations with IPF, and there is increasing evidence that microbiome composition also has a genetic basis. Previous studies have identified genetic signals involved in lung defence and cell proliferation to IPF risk and progression, but their effects on microbiome composition remain unclear. To uncover potential causal mechanisms linking the microbiome and lung fibrosis, we explored previously reported genetic association signals and their relationship with gut microbiome composition and IPF risk and outcomes. This revealed shared signals between gut microbiome variation and IPF that warrant further investigation. Our findings emphasise the value of further research leveraging genetic associations to improve our understanding of causal disease mechanisms and identify novel therapeutic opportunities.

## Background

Idiopathic pulmonary fibrosis (IPF) is a poorly understood chronic lung disease characterised by progressive fibrosis, with poor survival and limited treatment options. IPF is thought to result from an aberrant response to lung injury, culminating in an exaggerated healing response with excessive deposition of extracellular matrix in the interstitium (1) (2) (3). Genetic studies have emphasised high heritability and common polygenic aetiology in the development of IPF. There are more than 30 replicated common genetic signals associated with IPF (3) implicating genes involved in host defence, cell proliferation and signalling among others.

The lung harbours a diverse microbial community even in the absence of infection, and changes in its composition have been associated with disease progression in IPF (4) (5). Candidate gene studies of acute exacerbations-related death in IPF, identified that a specific genetic variant in *TLR3*, which plays a critical role in pathogen recognition and innate immunity activation, deregulates the lung microbiome in IPF and reduces lung fibroblast responses (6). This suggests that it may have a role in mortality associated with acute exacerbations induced by viruses and bacteria. In addition, the strongest genetic risk factor for IPF (the *MUC5B* promoter variant, rs35705950) was independently associated with lung bacterial burden in IPF patients (5). However, studies investigating the effects of host genetics on the airway or lung microbiome in health or disease are still needed, as previous studies have been limited in terms of sample size.

The gut microbiome has emerged as a promising area of investigation in respiratory diseases, due to its critical role in regulating immune responses and maintaining systemic homeostasis (7) (8) (9). Dysbiosis in gut microbiome composition can influence inflammation and immune modulation, potentially affecting disease progression. People with IPF have been shown to have altered gut microbiota profiles compared to healthy controls, characterised by decreased diversity and changes in the abundance of specific bacterial taxa (10). Moreover, animal studies support the role of the lung-gut axis in IPF pathogenesis, and suggest that altering the gut microbiota, through the use of antibiotics, probiotics, or faecal microbiota transplantation can moderate the severity of lung fibrosis (11) (12).

A study of the effect of host genetics on gut microbiome composition in the general population (N=18,340) identified 31 genetic loci influencing the microbiome at genome-wide significance (*p*<5×10^−8^) (13). Interestingly, one of the signals was located near the *FUT2* gene, which has been shown to affect exacerbation frequency and airway microbiota in patients with non-cystic fibrosis bronchiectasis (14). This signal has also been associated with lung function and chronic sputum production in the general population (15). Mendelian randomisation studies have shown evidence of potential causal relationships between specific gut bacterial taxa and IPF susceptibility and lung function, therefore supporting the importance of the gut microbiome in IPF and suggesting that interventions targeting microbiome-related pathways might have therapeutic benefit (16) (17) (18).

Our aim was to identify shared genetic signals and pathways involved in IPF pathogenesis and gut microbiome composition that could highlight specific causal links, warranting further research and informing the development of new therapeutic approaches. Given the limited availability of gut microbiome data from IPF patients, we took genetic associations with gut microbiome composition in the general population and assessed overlap with genetic associations with IPF susceptibility, progression (measured using lung function decline) and transplant-free survival times after IPF diagnosis.

## Methods & results

Of 31 genetic regions associated with the regulation of the gut microbiome (13), two signals were nominally (*p*<0.05) associated with IPF susceptibility (5,159 IPF cases and 27,459 controls) (3) and a further four were nominally associated with IPF disease progression (including lung capacity, FVC; and gas transfer, DLCO; N=1,048 individuals with IPF) (19). The most significant association was an intronic signal in the *NTRK2* gene, previously implicated in fibroblast activation in pulmonary fibrosis (20), which was associated with slower lung function decline in IPF (n=1,048, beta=31.2ml/year (95% CI: 7.5 - 54.9), *p*=6.62×10^−3^), and a decrease in the bacterial genus *Oxalobacter* in the general population (**Table 1a**).

**Table 1.**
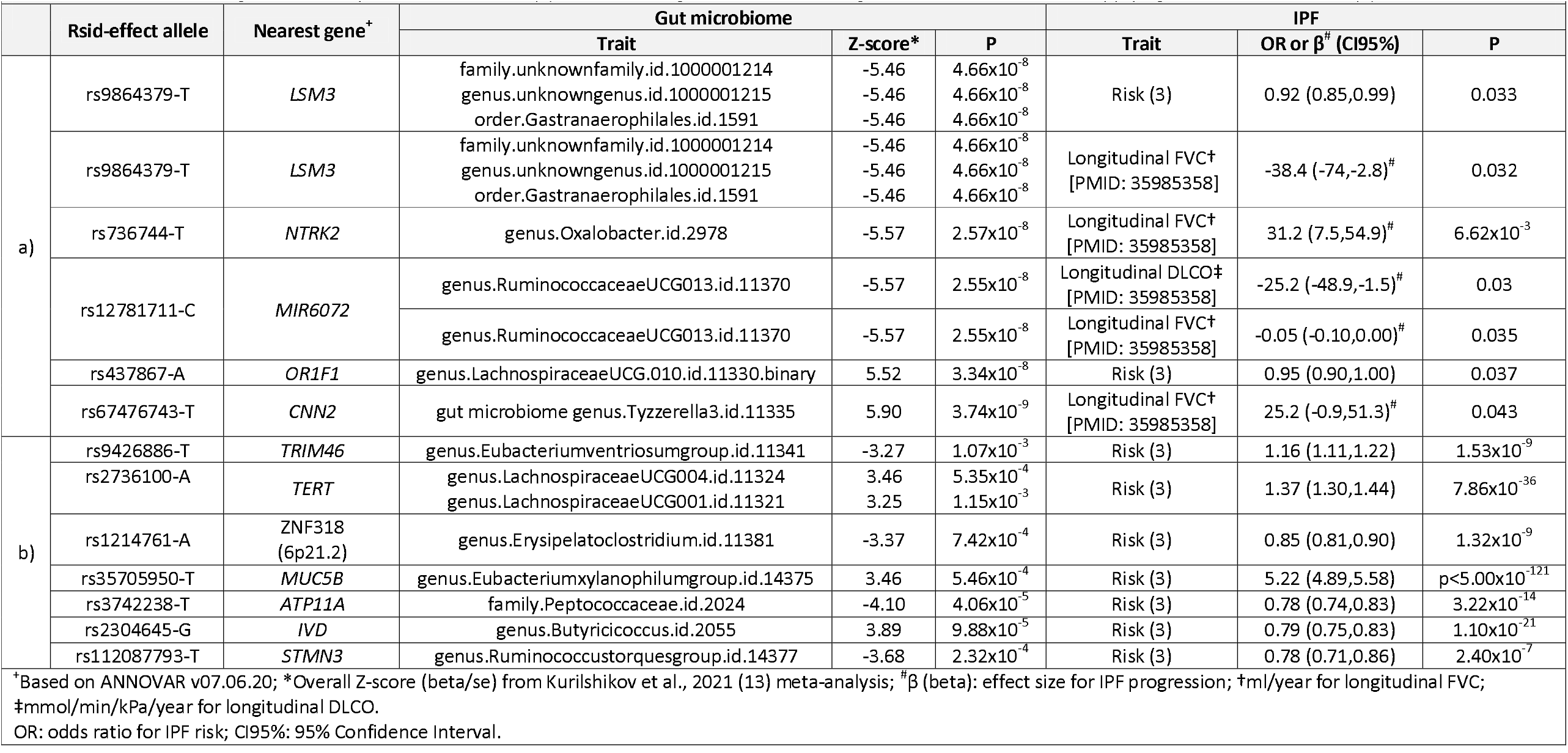
Gut microbiome signals nominally associated with IPF (a), and IPF risk signals associated with gut microbiome GWAS after applying Bonferroni correction (b).

Twenty-four of 35 previously reported association signals for IPF risk were at least nominally significant for association with at least one gut bacterial taxon. Seven signals were significant after accounting for 35 tests (*p*<1.4×10^−3^), although none would have passed a more stringent threshold of also accounting for 211 taxa; 7,385 tests, *p*<6.77×10^− 6^ (**Table 1b)**.

The most significant shared signal (p=4.06×10^−5^) was a single nucleotide polymorphism (SNP) at the *ATP11A* gene associated with decreased IPF risk (rs3742238-T) and with decreased relative abundance of *Peptococcaceae* family. The second most significant signal (*p*=9.88×10^−5^) was a SNP allele at the Isovaleryl-CoA Dehydrogenase (*IVD)* gene associated with decreased IPF risk (rs2304645-G) and with increased relative abundance of *Butyricicoccus* genus. To identify potential shared causal variants between gut microbiome variation and IPF, we performed colocalisation analysis using the coloc package (https://github.com/chr1swallace/coloc) in R. Both signals showed a high posterior probability (PP) of shared a causal variant contributing to IPF risk and *Peptococcaceae* (PP = 0.945) and *Butyricicoccus* (PP = 0.918) relative abundance. These taxa have not previously been linked to IPF.

## Discussion

Some treatments have been shown to modulate the gut microbiome in pulmonary fibrosis. For example, the administration of a traditional medicine used for cough and sputum production (amygdalus mongolica oil) in animal models increased intestinal diversity and the abundance of several bacterial taxa, including *Peptococcaceae*, which was associated with reduced oxidative and inflammatory damage and attenuation of fibrosis severity (9). Although this appears to contradict our finding of a genetic link between increased fibrosis risk and *Peptococcaceae* diversity, this may reflect context-dependent effects, whereby shifts in the microbiome associated with disease susceptibility differ from those observed during therapeutic modulation.

The *ATP11A* gene encodes an integral membrane ATPase involved in the innate immune response, with reduced protein levels linked to increased inflammation (21). The IVD enzyme plays a crucial role in breaking down the amino acid leucine within the mitochondria, an essential process for generating energy and preventing the accumulation of toxic by-products. Previous studies in inflammatory bowel disease have suggested that maintaining *Butyricicoccus* levels may reduce disease risk or slow progression (22). This raises the possibility that a similar intervention might have benefit for individuals with IPF or those at high risk of disease (23). Further investigation is needed to clarify the relationships between the genetic association signals and microbiome composition to establish whether they reflect causal effects or pleiotropy.

## Conclusion

In conclusion, our study leveraged existing data to identify shared genetic associations of gut microbiome composition and IPF that may indicate a causal role of the gut microbiome in the development of fibrotic lung disease. These findings require further validation to determine whether interventions targeting microbiome composition can meaningfully improve patient outcomes. Integrating host genetic data with microbiome profiles is essential to distinguish causal relationships from consequences of disease, thereby enabling the identification of interventions that may modify disease progression or even aid in prevention.

## Data

IPF GWAS data is available from: https://github.com/genomicsITER/PFgenetics. Gut microbiome GWAS in general population data is available from: www.mibiogen.org.

## Ethics

This study used publicly available summary statistics. No new data were generated and no individual-level data were used.

## Authors contribution

THB, LC, PLM and LVW designed the study. THB, RJA and OCL performed the analyses. THB and LVW wrote the first draft of the manuscript. All authors revised and approved the final version of the manuscript.

